# Rapid development of COVID-19 rapid diagnostics for low resource settings: accelerating delivery through transparency, responsiveness, and open collaboration

**DOI:** 10.1101/2020.04.29.20082099

**Authors:** Emily R Adams, Yolanda Augustin, Rachel L Byrne, David J Clark, Michael Cocozza, Ana I Cubas-Atienzar, Luis E. Cuevas, Martina Cusinato, Benedict M O Davies, Mark Davis, Paul Davis, Annelyse Duvoix, Nicholas M Eckersley, Thomas Edwards, Tom Fletcher, Alice J Fraser, Gala Garrod, Linda Hadcocks, Qinxue Hu, Michael Johnson, Grant A Kay, Katherine Keymer, Daniela Kirwan, Kesja Klekotko, Zawditu Lewis, Jenifer Mason, Josie Mensah-Kane, Stefanie Menzies, Irene Monahan, Catherine M Moore, Gerhard Nebe-von-Caron, Sophie I Owen, Tim Planche, Chris Sainter, James Schouten, Henry M Staines, Lance Turtle, Chris Williams, John Wilkins, Kevin Woolston, Amadou Alpha Sall, Joseph R A Fitchett, Sanjeev Krishna

**Author notes:** Corresponding authors: Sanjeev Krishna, Emily Adams.

## Abstract

Here we describe an open and transparent consortium for the rapid development of COVID-19 rapid diagnostics tests. We report diagnostic accuracy data on the Mologic manufactured IgG COVID-19 ELISA on known positive serum samples and on a panel of known negative respiratory and viral serum samples pre-December 2019.

In January, Mologic, embarked on a product development pathway for COVID-19 diagnostics focusing on ELISA and rapid diagnostic tests (RDTs), with anticipated funding from Wellcome Trust and DFID.

834 clinical samples from known COVID-19 patients and hospital negative controls were tested on Mologic’s IgG ELISA. The reported sensitivity on 270 clinical samples from 124 prospectively enrolled patients was 94% (95% CI: 89.60% - 96.81%) on day 10 or more post laboratory diagnosis, and 96% (95% CI: 84.85% - 99.46%) between 14–21 days post symptom onset. A specificity panel comprising 564 samples collected pre-December 2019 were tested to include most common respiratory pathogens, other types of coronavirus, and flaviviruses. Specificity in this panel was 97% (95% CI: 95.65% - 98.50%).

This is the first in a series of Mologic products for COVID-19, which will be deployed for COVID-19 diagnosis, contact tracing and sero-epidemiological studies to estimate disease burden and transmission with a focus on ensuring access, affordability, and availability to low-resource settings.

## Introduction

On the 31st of January, six weeks after the first report in Wuhan, China, of SARS-CoV-2, the causative pathogen of Coronavirus Disease 2019 (COVID-19), and 24 hours after the World Health Organization (WHO) declared COVID-19 a public health emergency, the United Kingdom reported its first two imported cases. By the end of April >20,000 hospitalized patients with COVID-19 had died in the UK and >200,000 fatalities have occurred worldwide, with the pandemic spreading to more than 190 countries^1^. The WHO advises countries to ‘test, test, test’ as one of the pillars for COVID-19 control, although most countries don’t have enough tests for widespread deployment and testing strategies focus on identifying symptomatic patients who need hospitalization or for isolation and use among key workers.^2^

Current testing is almost exclusively reliant on RT-PCR to detect the virus RNA. These assays are sensitive and specific for SARS-CoV-2, but the scale of the COVID-19 pandemic has caused global shortages of essential reagents for testing, including those for sample collection, RNA extraction and pathogen detection. These have hampered the urgent scale-up of testing required, and has led to the politicization of diagnostic testing with reports of stockpiling resources.^3–5^ Nucleic acid amplification and detection technologies are generally not deployable at the point-of-need, as most of them require significant laboratory infrastructure, expertise, and skilled staff with appropriate personal protective equipment (PPE).

Governments and industry are increasing testing capacity through the accelerated development of simpler, less elaborate and more affordable rapid diagnostic tests (RDTs). These tests may detect viral protein antigens in nasopharyngeal swabs or oral fluid to determine acute infection, or antibodies in blood to indicate seroconversion after infection, similar to laboratory-based enzyme-linked immunosorbent assay (ELISA) technology. RDTs are more suitable for use at the point-of-need, because although they may offer lower sensitivity and specificity than RT-PCR-based tests in some circumstances, their ease of use and rapid readout of results make them important components of diagnostic and contact tracing control strategies, especially as tools for rapid triage at the frontline of hospitals, primary care facilities, and for reaching into the community and home. Antibody detection will also be key to informing exit strategies for the expected long tail of the pandemic and in supporting surveillance by identifying clusters of infection and convalescent cases or people with milder disease that could seed the infection to generate severe cases.^6^

The rollout of RDTs is currently marred by a lack of performance data, reference materials, numbers tested and transparency on how tests are being evaluated. Many RDTs have been rushed to market with limited or no independent evaluations of performance. Marketing information is typically based on internal evaluations with poorly designed studies with limited participant characterization. Governments have cancelled orders once independent evaluations have failed to replicate the claims of the manufacturer.^7^

We describe here a partnership set up to develop low-cost COVID-19 tests to detect (a) SARS-CoV-2 antigens and (b) circulating IgG, IgA and IgM antibodies using lateral flow assays (LFA) delivering results in <15 minutes and (c) complementary ELISA assays for laboratory-based mass screening for seroconversion. Development of these diagnostic tools is a result of a partnership between Mologic (UK), a biotechnology company committed to developing tests for low resource settings at cost of goods in line with Global Access Policy^8^, the Liverpool School of Tropical Medicine (LSTM, UK), St George’s University of London (SGUL, UK), and the Institut Pasteur de Dakar (IPD, Senegal). Our consortium includes a globally distributed testing network across four continents to evaluate novel RDTs in Malaysia, Kenya, Malawi, China, Spain, North America, and Latin America.

Our aim is to develop COVID-19 RDTs to meet the WHO provisional target product profiles (TPP) for point-of-need tests for antigen and antibody assays^9^. Antigen assays, could be used for the rapid triage of patients with presumptive COVID-19, to detect asymptomatic or presymptomatic carriers and mild infections for isolation^10^. Asymptomatic and subclinical infections play an important role in the transmission of SARS-CoV-2 to close contacts, especially among cohabiting family members^11–13^ and may be responsible for 50%-80% of infections^14,15^, although, critically, we need accurate tests to confirm this hypothesis.

Furthermore, these point-of-need antigen tests will be formatted as self-tests, offering the additional benefit of being widely used in the home and community settings by professional testers for rapid triage, to test contacts of confirmed cases, and to decentralise surveillance. Antibody assays have been proposed as part of the ‘exit strategy’ from large scale lockdown of countries as a “back to work” test. However, RDTs remain insufficiently accurate due to their low positive predictive and negative value among low prevalence settings in the community, and because the presence of antibodies have not yet been shown to correlate with long-lasting immunity to reinfection.

## Methods

Following the declaration of COVID-19 as a Public Health Emergency of International Concern on 30^th^ January 2020^16^, an application by Mologic, LSTM and SGUL to develop and manufacture rapid tests for COVID-19 was considered successful for funding from the Wellcome Trust and the Department for International Development (DFID) on the 28^th^ of February. The program started on March 6^th^ and the first batch of 15 different prototypes arrived at LSTM and SGUL ready for evaluation two weeks from commencing, and 8 weeks ahead of the official award letter. The first batch of prototypes was evaluated against positive serum obtained from convalescent COVID-19 originally confirmed by Public Health England’s RT-PCR tests and provided by International Severe Acute Respiratory and Emerging Infection Consortium 4C (ISARIC) in the UK, and prospectively collected SGUL in-patient samples. Data from testing were shared and discussed across all partners on the 26^th^ of March. Second, third and fourth rounds of independent iterative validation across three sites were conducted in sequential weeks allowing Mologic to accelerate the best performing prototypes and optimise assay components.

We envisaged this approach that combined the strengths of industry and academia, would speed up the development of these critically needed tests. The subsequent robust and transparent evaluation will provide all stakeholders with accurate information, which is critical for the informed decision of test implementation and their correct placement in the UK’s testing strategy. Prototype RDTs and the ELISA have been evaluated in prospectively collected excess diagnostic waste clinical samples of patients with RT-PCR confirmed COVID-19 at SGUL and a specificity panel from LSTM, Mologic and SGUL of pre-December 2019 bio-banked serum samples.

At SGUL, anonymised excess diagnostic material (and relevant clinical data) from patient samples tested for SARS-CoV-2 in South West London Pathology (SWLP) microbiology laboratory at St George’s, University Hospitals NHS Foundation Trust (SGHFT) were collected. RT-PCR confirmation of SARS-CoV-2 infection in patients from which excess diagnostic material were collected used nasopharyngeal swabs placed into Sigma Virocult®. An aliquot was subsequently placed into Roche PCR lysis buffer for RNA extraction using Roche RNA extraction kits on a Magnapure (with extracts placed into 96 well plates). SARS-CoV-2 detection was undertaken using the Altona Diagnostics RealStar®SARS-CoV-2 RT-PCR Kit (S and E genes).

We have also engaged with the ISARIC 4C consortium, who are providing 50 serum samples on an ongoing basis samples for testing, and we will publish results of these rounds of testing once we reach the sample size.

## Ethics

Clinical sample collection was approved at SGUL as part of the DARTS study - IRAS project ID: 282104; REC reference: 20/SC/0171

## Results

Here we provide validation data for a novel IgG ELISA for SARS-CoV-2. These data were generated using 96-well plate kits provided by Mologic, in which an early generation assay used NP, S1 and S2 antigens and the finalized version used only NP and S2 antigens (with S1 being removed). Assays were undertaken following the manufacturer’s protocol, initially in triplicate and subsequently in duplicate, with each plate having a diluent (negative), cut off and positive controls (results were considered positive if they were 10% above the cut off value). Data are presented in Tables 1 and 2.

**Table 1.** Performance Evaluation Data. A total of 270 positive samples (laboratory confirmed SARS-CoV-2 by RT-PCR, ranging from day 0 to day 42) and 564 negative samples (from 2018–2019) were tested, with the following results.

| **834 clinical samples analysed** | **% (N)** | **95% CI** |
| --- | --- | --- |
| **Sensitivity**<br>Total SARS-CoV-2 positive samples ranging from day 0 - 42 post laboratory diagnosis with RT-PCR | 88% (238/270) | 83.68% - 91.75% |
| **Specificity**<br>Total SARS-CoV-2 negative samples ranging from 2018-2019 | 97% (549/564) | 95.65% - 98.50% |

**Table 2.** A) Sensitivity stratified by days post swab for RT-PCR.

| **Days post laboratory diagnosis** | **Sensitivity (%)** | **95% CI** |
| --- | --- | --- |
| $\leq 7$ | 70% (37/53) | 55.66% - 81.66% |
| $\geq 8$ | 93% (199/215) | 88.20% - 95.69% |
| $\geq 10$ | 94% (185/197) | 89.60% - 96.81% |
| $\geq 14$ | 94% (150/159) | 89.53% - 97.38% |

**Table 2.**
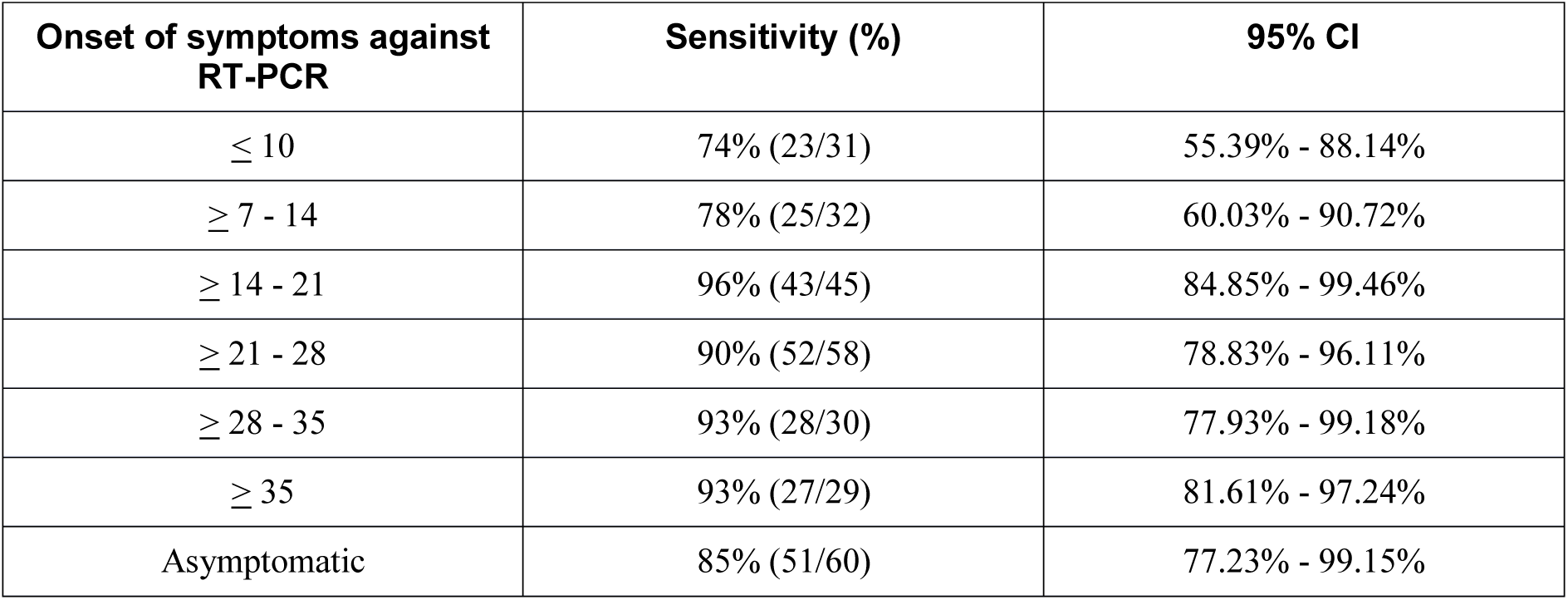
B) Sensitivity stratified by onset of symptoms.

| **Onset of symptoms against RT-PCR** | **Sensitivity (%)** | **95% CI** |
| --- | --- | --- |
| $\leq 10$ | 74% (23/31) | 55.39% - 88.14% |
| $\geq 7 - 14$ | 78% (25/32) | 60.03% - 90.72% |
| $\geq 14 - 21$ | 96% (43/45) | 84.85% - 99.46% |
| $\geq 21 - 28$ | 90% (52/58) | 78.83% - 96.11% |
| $\geq 28 - 35$ | 93% (28/30) | 77.93% - 99.18% |
| $\geq 35$ | 93% (27/29) | 81.61% - 97.24% |
| Asymptomatic | 85% (51/60) | 77.23% - 99.15% |

**Table 3.**
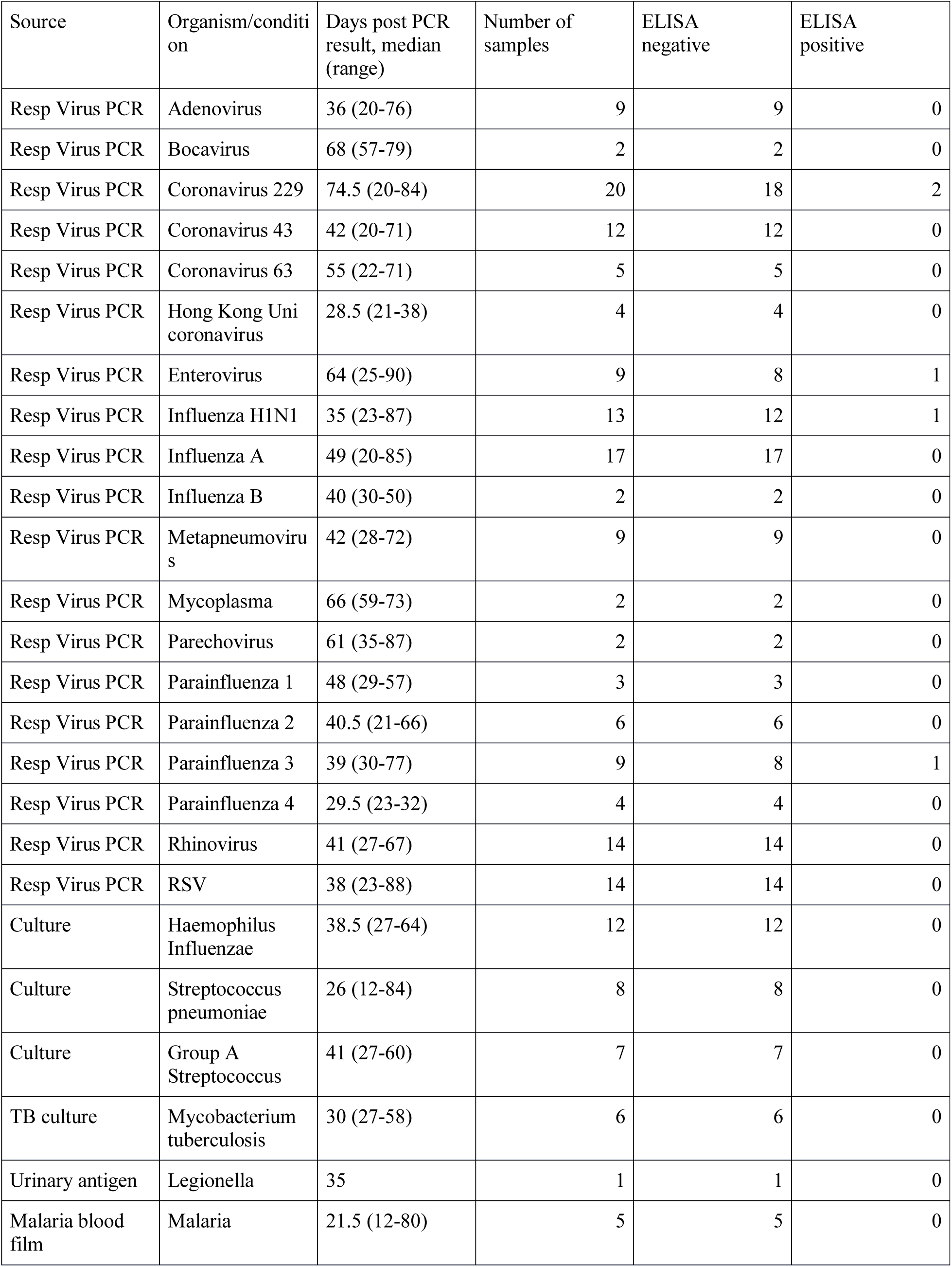
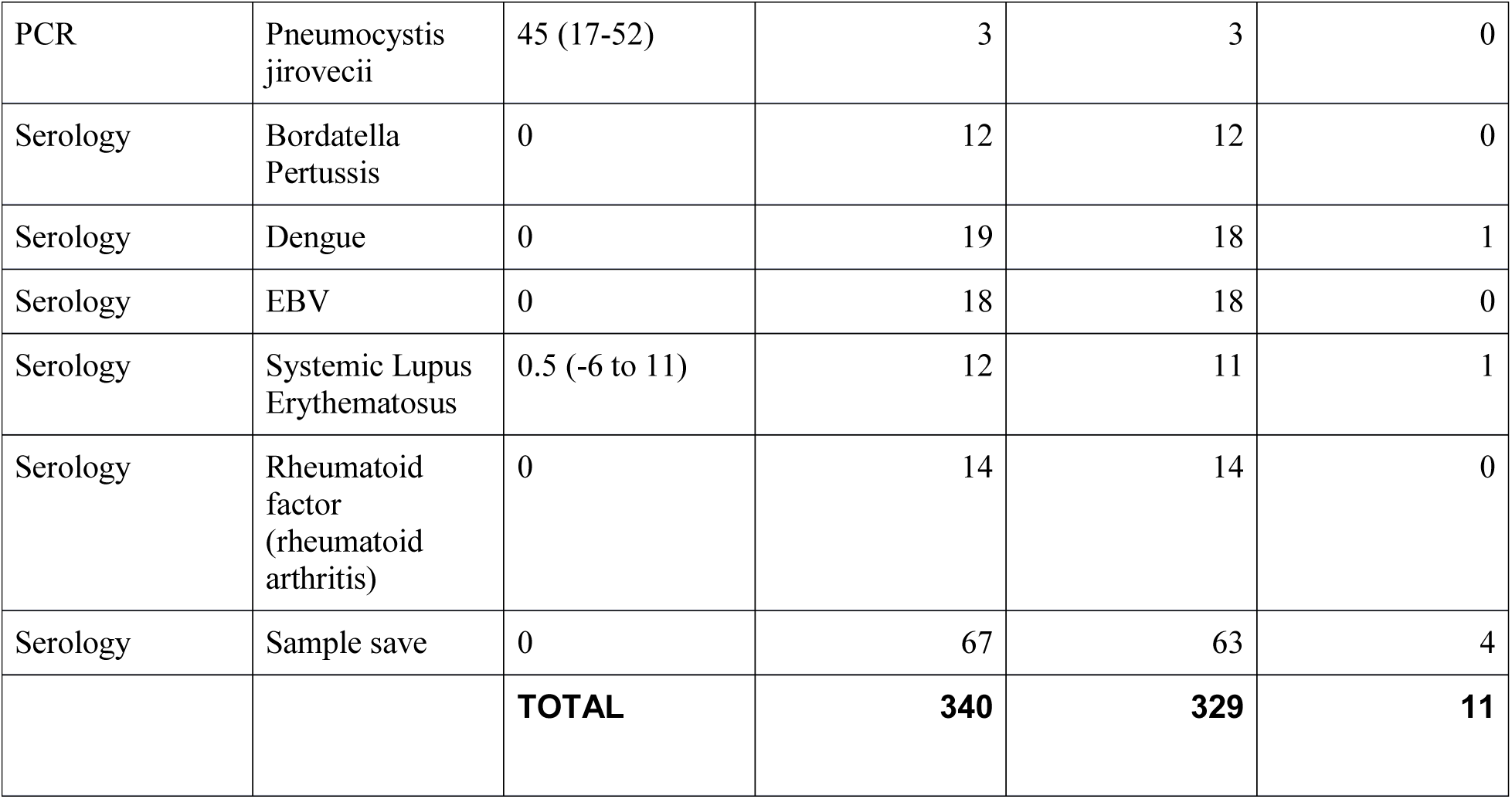
A) Cross reactivity: SARS-CoV-2 negative samples that are positive for a range of other viruses.

A total of 270 positive samples laboratory confirmed SARS-CoV-2 by RT-PCR, ranging from day 0 to day 42, and 564 negative samples (from 2018–2019) were tested. Clinical samples were analysed from a total of 124 patients, including from 50 women (40.3%) and 74 men (59.7%) with an age range of 26 to 88 years. Overall, the sensitivity was 88% (95% CI: 83.68% - 91.75%) in all samples (238 of 270 positive samples). The specificity was 97% (95% CI: 95.65% - 98.50%) (549 of 564 samples tested).

Sensitivity of the IgG ELISA was 94% (95% CI: 89.60% - 96.81%; 185 of 197 positive samples) among samples greater than 10 from initial laboratory diagnosis. Sensitivity among symptomatic cases was 96% between 14 and 21 days of symptom onset (95% CI: 84.85% - 99.46%) compared with 78% among cases less than 14 days from symptom onset (95% CI: 64.04% - 88.47%).

A total of 209 samples (77.4%) were collected from 90 individuals reporting symptoms of COVID-19 and 61 samples (22.6%) from 34 individuals who were asymptomatic at first swab collection. Of the 34 individuals who were asymptomatic at presentation and positive by RT-PCR, 29 were also COVID-19 positive with IgG ELISA (85.3%). Of the remaining 5 individuals, 3 seroconverted (8.8% of asymptomatic cases on presentation) on repeat testing and 2 individuals remained negative on the IgG ELISA despite repeat testing (5.8% of asymptomatic cases on presentation). Among the 90 RT-PCR positive symptomatic individuals, 11 were IgG ELISA negative at presentation (12.2%), 6 seroconverted on repeat testing (6.7% of symptomatic cases on presentation)., and 5 individuals remained negative on IgG ELISA (5.5% of symptomatic cases on presentation).

Time from swab to laboratory confirmation was comparable between ELISA positive and ELISA negative samples. However, ELISA negative samples were on average 3 days sooner to symptom onset than ELISA positive samples (20.5 days versus 23.5 days, respectively). In addition, swab collection was 6 days earlier among ELISA negative samples than ELISA positive samples (11.6 days versus 17.5 days, respectively) and laboratory confirmation was 5 days earlier (10.1 days versus 15.7 days, respectively). Collectively this suggests a proportion of ELISA negative samples are yet to manifest as positive for IgG detection.

## Discussion

### Challenges of RDT development

The development of RDTs takes longer than that of RT-PCR tests, which were designed and developed within days of the publication of the first viral genomes^17–19^, and then validated within weeks^20,21^. RDTs require the production of viral protein antigen, in the case of antibody detection, or the production of specific antibodies in the case of antigen detection.

Rapid tests are difficult to develop without access to well-characterised patient samples, unlike RT-PCR tests which can be developed using synthetic RNA with the viral target sequence. However, not all RDTs are created equal. In existing RDT markets there is a substantial variation of performance and, for example, the sensitivity of malaria RDTs varies between 31.7% and 100% across manufacturers^22^. The same variations are documented for Dengue^23^ and leishmaniasis^24^, to name but a few diseases. It is thus essential that new COVID-19 tests are subjected to robust independent evaluations, and that results from evaluations are transparent and freely available for scrutiny by governments and the public. The methodology of an evaluation, including the study population, the reference test used, and the handling of the sample have a large bearing on the results of the study. A badly designed study will either under- or overestimate a test’s accuracy, invalidating the decision of whether to implement that particular test or not.

### Transparency and Openness

In the UK, the target sensitivity and specificity from the MHRA was published on the 8^th^ April 2020, several days after ELISAs and RDTs were reported to fail evaluations as tests for ‘immunity passports’ and seroprevalence studies.^25^ The required accuracy for such tests is currently specified at >98% sensitivity and specificity (update 24^th^ April 2020^26^). However, all manufacturers reported lower accuracy levels than this threshold from their own evaluations, and therefore would not have expected to pass these stringent criteria. The current lack of transparency and rapid feedback to product developers removes the opportunity for evaluation data to inform test optimisation and to develop improved products. Additionally, company names were not released with the evaluation data due to non-disclosure agreements^25^. Not having a TPP by WHO for COVID-19 diagnostics – 3 months since the declaration of a global public health emergency only serves to hamper the accelerated development of novel technologies to support pandemic control. Release of preliminary data to the scientific community for consultation and scrutiny would allow for a wider discussion of results, and a greater understanding of likely test accuracy. Furthermore, we would encourage evaluations within networks in the UK; the greater the sample number, the more accurate predictions are, whereas current MHRA criteria may perversely encourage small, poorly designed studies with large confidence intervals to hit the targets.

As a team of academic evaluators, at the LSTM and St George’s, University of London, and Mologic, a biotechnology company and manufacturer of rapid diagnostic tests, we have sought to aid the development of new RDTs for SARS-CoV-2 antigens and antibodies. We have formed an accelerated validation partnership enabling the independent evaluation of prototype tests, with continual feedback to the test developer allowing for iterative optimisation of the assay. During this process we strive to make results available to the scientific community as rapidly as possible, both via publication in journals, and in frequently updated results summaries on public domains such as BioRxiv (https://www.biorxiv.org) and MedRxiv (https://www.medrxiv.org). This is our first submission of the ELISA IgG data which looks extremely promising and is now available from Mologic with increased manufacturing capability aided by DFID, FIND and BMGF.

## Data Availability

All raw data available

## Funding and Acknowledgements

DFID/Wellcome Epidemic Preparedness coronavirus, 220764/Z/20/Z.

ERA, LEC, LT and TF are affiliated to the National Institute for Health Research Health Protection Research Unit (NIHR HPRU) in Emerging and Zoonotic Infections (NIHR200907) at University of Liverpool in partnership with Public Health England (PHE), in collaboration with Liverpool School of Tropical Medicine and the University of Oxford. The views expressed are those of the author(s) and not necessarily those of the NHS, the NIHR, the Department of Health or Public Health England.

HMS is supported by the Wellcome Trust Institutional Strategic Support Fund (204809/Z/16/Z) awarded to St. George’s University of London.

LT is supported by the Wellcome Trust (grant number 205228/Z/16/Z) We wish to thank the National Association of Blood Bikes (NABB) for their unwavering assistance in transporting samples, prototypes, and validated devices between Liverpool, Bedford, and London. Without the NABB, we could not have delivered two CE marked products in 8 weeks from project launch.

## Authors’ contributions

ERA, JRAF, HMS, LEC, SK, TP conceived study.

ACA, AJF, GAK, SIO, RLB, GG DJC, BMOD, NME, LH, QH, IM, CMM, HMS, SK performed technical work and handled clinical samples.

JM, KK, LT & TF provided clinical data for samples at LSTM.

YA, MC, DK, SK, TP & HMS developed the DARTS protocol and provided clinical data for samples at SGUL

MC, MD, PD, AD, AJ, KK, ZL, JMK, GNvC, CS, JC, JW, KW, AAS, JRAF developed the ELISA test at Mologic.

ERA, ACA, TE, LEC, SK wrote the first draft of the article.

All authors reviewed the manuscript before submission.

## Tables and Figures

### Performance Characteristics

**Figure 1:**
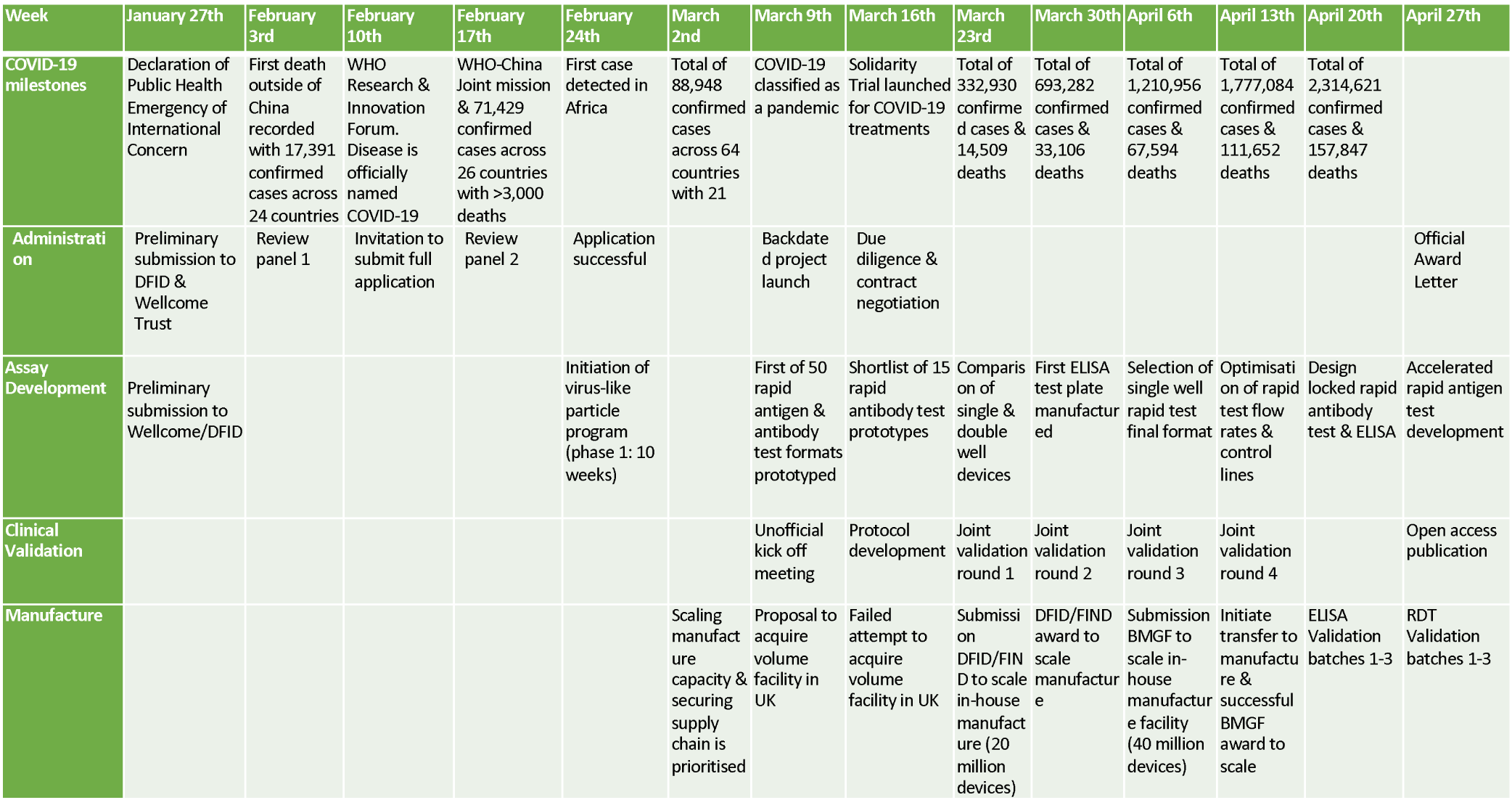
Timeline of events for Mologic IgG ELISA, including COVID-19 milestones, Administration, Assay development, Clinical Validation and Manufacture.

**Figure 2.**
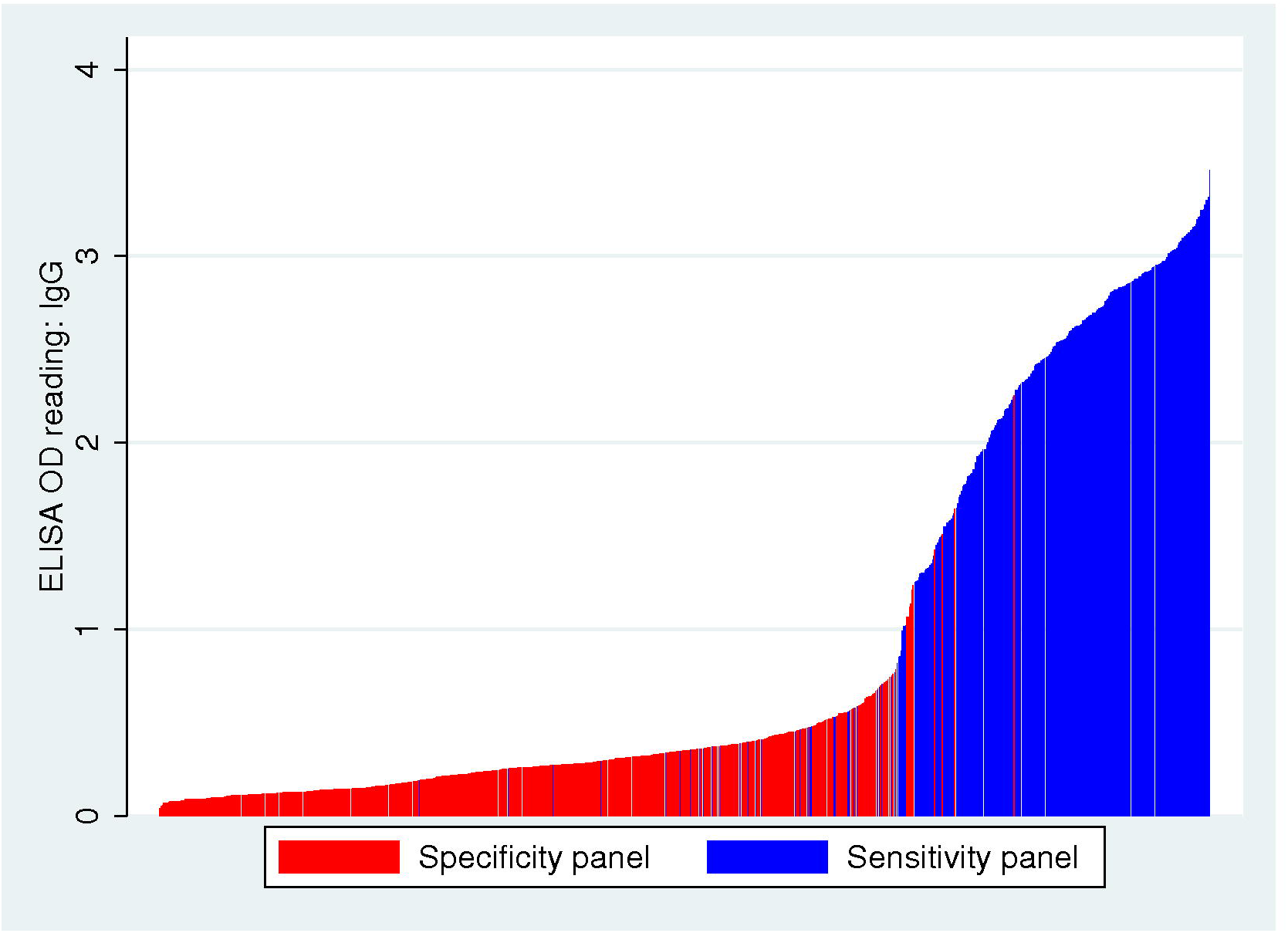
Distribution of OD readout from the IgG ELISA among 270 laboratory confirmed samples of COVID-19 from 124 patients and 564 negative samples.

**Figure 3.**
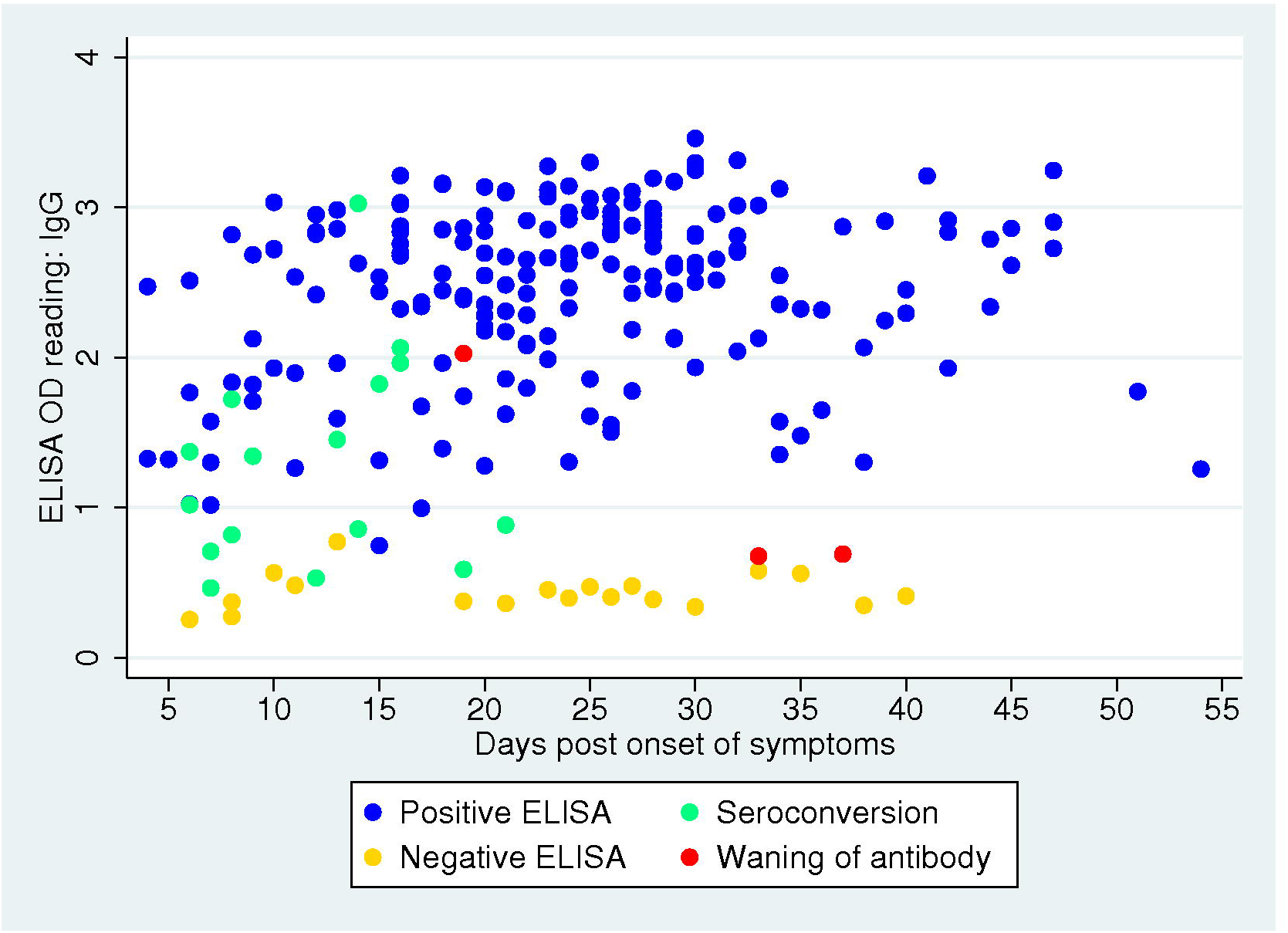
Distribution of OD readout from the IgG ELISA by seroconversion status among symptomatic patient only. Blue represents Positive ELISA (n=77 patients with 225 samples), yellow represents Negative ELISA (n=7 patients with 21 samples), green represents Seroconversion from negative to positive ELISA on repeat testing (n=8 patients with 21 samples), and red represents Waning of immune response from positive to negative ELISA (n=1 with 3 samples).

**Figure 4.**
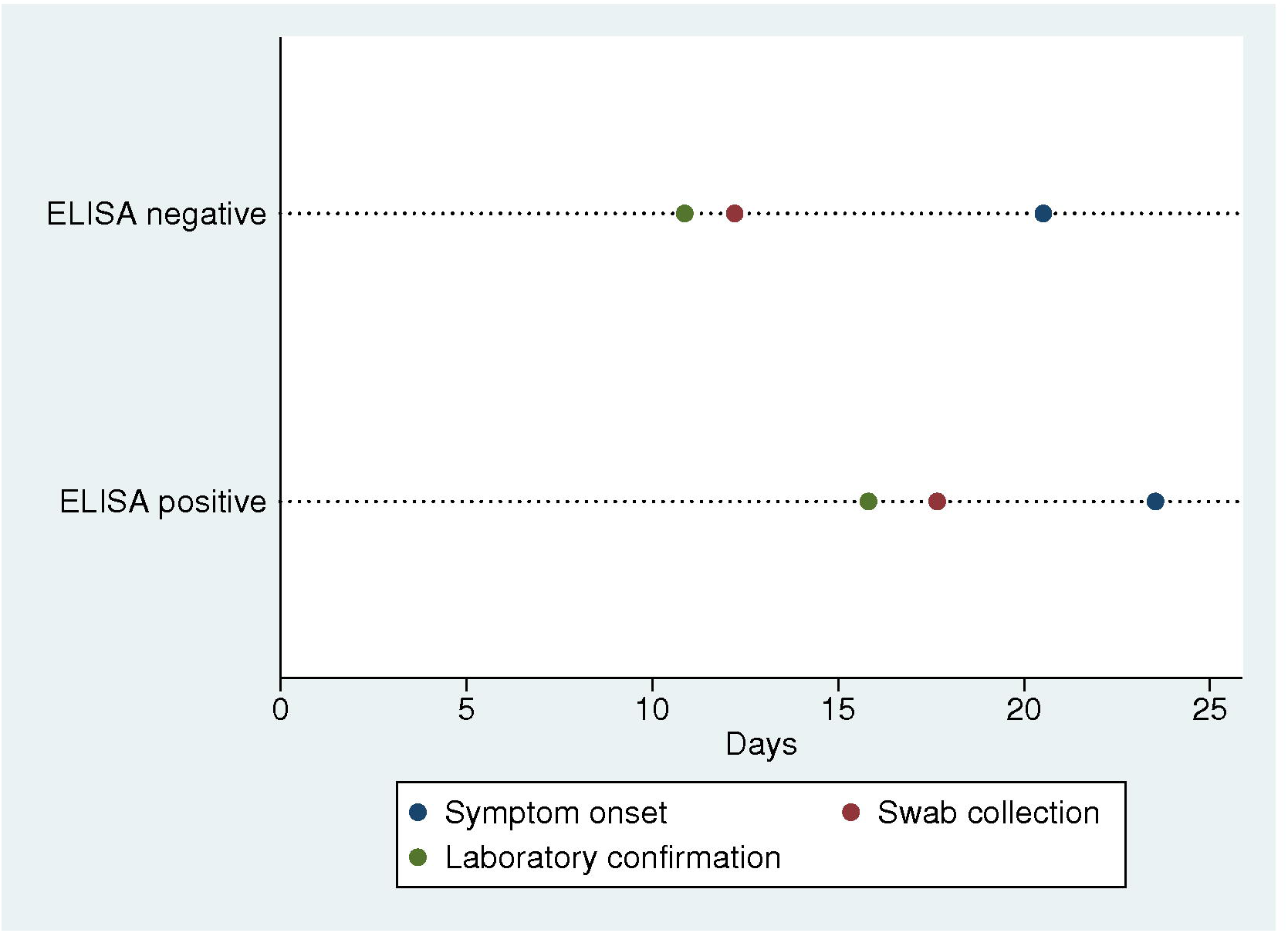
Comparison in days of timelines from symptom onset, swab processing, and laboratory confirmation among ELISA positive and ELISA negative samples.

## References

1 World Health Organization. Novel Coronavirus (2019-nCoV) Situation Report - 97. WHO Bull. 2020. https://www.who.int/docs/default-source/coronaviruse/situation-reports/20200426-sitrep-97-covid-19.pdf (accessed April 6, 2020).

2 World Health Organization. Laboratory testing for 2019 novel coronavirus (2019-nCoV) in suspected human cases. WHO - Interim Guid 2020. https://apps.who.int/iris/bitstream/handle/10665/330676/9789240000971-eng.pdf (accessed April 6, 2020).

3 Knaus C. Australian health department says supply of key component in coronavirus testing under strain. Guard. 2020. https://www.theguardian.com/world/2020/mar/16/australian-doctors-warn-coronavirus-testing-compromised-by-failure-to-stockpile-key-chemical-reagent (accessed April 6, 2020).

4 Kuznia R, Curt D, Griffin D. Severe shortages of swabs and other supplies hamper coronavirus testing - CNN. CNN US. https://www.cnn.com/2020/03/18/us/coronovirus-testing-supply-shortages-invs/index.html (accessed April 6, 2020).

5 Akst J. RNA Extraction Kits for COVID-19 Tests Are in Short Supply in US. Sci. 2020. https://www.the-scientist.com/news-opinion/rna-extraction-kits-for-covid-19-tests-are-in-short-supply-in-us-67250 (accessed April 6, 2020).

6 Yong SE, Anderson DE, Wei WE, et al. Connecting clusters of COVID-19: an epidemiological and serological investigation. Lancet Infect Dis. 2020; DOI: 10.1016/S1473-3099(20)30273-5.

7 Jones S. Coronavirus test kits withdrawn in Spain over poor accuracy rate. Guard. 2020. https://www.theguardian.com/world/2020/mar/27/coronavirus-test-kits-withdrawn-spain-poor-accuracy-rate (accessed April 26, 2020).

8 Bill and Melinda Gates foundation. Global access in action. http://globalaccess.gatesfoundation.org (accessed April 26, 2020).

9 World Health Organization. Instructions for Submission Requirements: In vitro diagnostics (IVDs) Detecting Antibodies to SARS-CoV-2 virus. WHO-Prequalification of In Vitro Diagnostics. 2020. https://www.who.int/diagnostics_laboratory/200417_pqt_ivd_eul_requirements_ncov_rdt_antibody.pdf (accessed April 26, 2020).

10 Aguirre-Duarte N. Can people with asymptomatic or pre-symptomatic COVID-19 infect others: a systematic review of primary data. MedRxiv. 2020; DOI: 10.1101/2020.04.08.20054023

11 Chan JFW, Yuan S, Kok KH, et al. A familial cluster of pneumonia associated with the 2019 novel coronavirus indicating person-to-person transmission: a study of a family cluster. Lancet. 2020; DOI:10.1016/S0140-6736(20)30154-9.

12 Phan LT, Nguyen TV., Luong QC, et al. Importation and human-to-human transmission of a novel coronavirus in Vietnam. N Engl J Med. 2020; DOI:10.1056/NEJMc2001272.

13 Wu JT, Leung K, Leung GM. Nowcasting and forecasting the potential domestic and international spread of the 2019-nCoV outbreak originating in Wuhan, China: a modelling study. Lancet. 2020; DOI:10.1016/S0140-6736(20)30260-9.

14 Li R, Pei S, Chen B, et al. Substantial undocumented infection facilitates the rapid dissemination of novel coronavirus (COVID-19). MedRxiv. 2020; DOI:10.1101/2020.02.14.20023127.

15 Arons MM, Hatfield KM, Reddy SC et al. Presymptomatic SARS-CoV-2 infections and transmissions in a skilled nursing facility. N Engl J Med. 2020; DOI:10.1056/NEJMoa2008457

16 World Health Organization. Coronavirus disease 2019 (COVID-19) Situation Report-68. WHO 2020. https://www.who.int/docs/default-source/coronaviruse/situation-reports/20200328-sitrep-68-covid-19.pdf (accessed April 26, 2020).

17 World Health Organization (WHO). National laboratories, Coronavirus disease (COVID-19) technical guidance: Laboratory testing for 2019-nCoV in humans. 2020. https://www.who.int/emergencies/diseases/novel-coronavirus-2019/technical-guidance/laboratory-guidance (accessed April 7, 2020).

18 Centers for Disease Control and Prevention (CDC). Coronavirus disease (Covid-19). Information for laboratories, Real-Time PCR resources. 2020. https://www.cdc.gov/coronavirus/2019-ncov/lab/rt-pcr-panel-primer-probes.html (accessed April 6, 2020)

19 Corman V, Bleicker T, Bru□nink S et al. Diagnostic detection of 2019-nCoV by real-time RT-PCR. 2020 https://www.who.int/docs/default-source/coronaviruse/protocol-v2-1.pdf (accessed April 6, 2020)

20 Jung YJ, Park G-S, Moon JH, et al. Comparative analysis of primer-probe sets for the laboratory confirmation of SARS-CoV-2. BioRxiv. 2020; DOI:10.1101/2020.02.25.964775.

21 Corman VM, Landt O, Kaiser M, et al. Detection of 2019 novel coronavirus (2019-nCoV) by real-time RT-PCR. Euro Surveill. 2020; DOI:10.2807/1560-7917.ES.2020.25.3.2000045.

22 World Health Organization. Malaria Rapid Diagnostic Test Performance. WHO 2009. https://www.who.int/malaria/publications/atoz/9789241510035/en/(accessed April 7, 2020).

23 Murtagh M. Dengue Virus Infection Diagnostics Landscape. WHO 2017. http://www.idc-dx.org/resources/dengue-virus-infection-diagnostics-landscape (accessed April 26, 2020).

24 Adams, E., Hasker, E. and Cunningham, J. Visceral leishmaniasis rapid diagnostic test performance.2011 on behalf of WHO-TDR.WHO. https://www.who.int/tdr/publications/documents/vl-rdt-evaluation.pdf (accessed April 27, 2020)

25 Adams ER, Anand R, Andersson MI, et al. Evaluation of antibody testing for SARS-Cov-2 using ELISA and lateral flow immunoassays. MedRxiv. 2020; DOI:10.1101/2020.04.15.20066407.

26 GOV.UK. Coronavirus (COVID-19): What do you need to know. Medical device regulation and safety. https://www.gov.uk/topic/medicines-medical-devices-blood/medical-devices-regulation-safety (accessed April 27, 2020).

